# COVID-19 Fatality and Comorbidity Risk Factors among Diagnosed Patients in Mexico

**DOI:** 10.1101/2020.04.21.20074591

**Authors:** Patricio Solís, Hiram Carreño

## Abstract

As of April 18, 2020, 2.16 million patients in the world had been tested positive with Coronavirus (COVID-19) and 146,088 had died, which accounts for a case fatality rate of 6.76%. In Mexico, according to official statistics (April 18), 7,497 cases have been confirmed with 650 deaths, for a case fatality rate of 8.67%. These estimates, however, may not reflect the final fatality risk among COVID-19 confirmed patients, because they are based on cross-sectional counts of diagnosed and deceased patients, and therefore are not adjusted by time of exposure and right-censorship. In this paper we estimate fatality risks based on survival analysis methods, calculated from individual-level data on symptomatic patients confirmed with COVID-19 recently released by the Mexican Ministry of Health. The estimated fatality risk after 35 days of onset of symptoms is 12.38% (95% CI: 11.37-13.47). Fatality risks sharply rise with age, and significantly increase for males (59%) and individuals with comorbidities (38%-168%, depending on the disease). Two reasons may explain the high COVID-19 related fatality risk observed in Mexico, despite its younger age structure: the high selectivity and self-selectivity in testing and the high prevalence of chronic-degenerative diseases.

## Introduction

Two important characteristics that differentiate the current COVID-19 pandemics from recent previous outbreaks such as SARS coronavirus and H1N1 influenza are its high rate of transmissibility and its apparently higher fatality rate (Liu et al 2020, Surveillances 2020). The rapid spread of the new infection represents a challenge for the timely calculation of epidemiological and demographic parameters. One of these challenges is the estimation of fatality risks.

As of April 18, 2020, 2.16 million patients in the world had been tested positive with Coronavirus (COVID-19) and 146,088 had died, which accounts for a fatality of 6.76%. In Mexico, according to official statistics (April 18), 7,497 cases have been confirmed with 650 deaths, for a fatality of 8.67% (Secretaría de Salud, 2020). However, these estimates may not reflect the final fatality rates among COVID-19 confirmed patients. Given that they are obtained from cumulative daily counts of confirmed patients and related deaths, they do not take into consideration the different exposure times of current confirmed patients, and therefore may be biased by right-censorship, particularly in countries in the early stages of evolution of the epidemics, such as Mexico, which accumulate a large proportion of recent cases.

In order to obtain unbiased estimates of fatality risk, it is necessary to consider patients’ time of exposure from the onset of symptoms and correct the biases in exposure associated to right-censorship. This may be achieved through the usual methods of survival analysis. These methods, however, require individual-level data with information about the date of the onset of symptoms, the date of death for the deceased, and basic demographic information.

With the exception of a handful of clinical studies (Shi et al. 2020; Zha et al. 2020; Zhou et al. 2020) data with such characteristics are unavailable at this moment for most countries. In the case of Mexico, the federal Ministry of Health (Secretaría de Salud) recently made public individual-level data of patients attended in medical units across the country who tested positive for COVID-19. This information, which is updated on a daily basis, allows us to estimate fatality risks for COVID-19 patients using survival analysis, as well as the possible effects of basic demographic factors, the presence of comorbidities and the type of medical institution on the risk of death.

## Data and Methods

Our analysis is based on 7,497 patients who tested positive for COVID-19 in Mexico, a subset of a larger dataset of 49,167 patients with COVID-19 related symptoms who sought attention in medical units and were tested with positive, negative (29,301 cases) and pending (12,369 cases) results. The information about these patients is reported on a daily basis to the Secretaría de Salud, which makes the integrated data available to the public and updated daily. We use the data set released on April 18, 2020^1^.

A first relevant characteristic of this dataset is the inclusion of the dates at the onset of symptoms and death, which has occurred in 650 cases. Basic demographic traits such as age and sex of the patient are also available. The data include information as well on pre-existing comorbidities such as high blood pressure, diabetes, chronic obstructive pulmonary disease (COPD), asthma, immunosuppression, cardiovascular diseases, obesity, chronic kidney disease (CKD) and smoking.

Medical attention in Mexico is highly segmented according to the type of medical institution. The data include information on the type of institution where the patient received attention. In addition, there is an identifier indicating whether the medical unit is a monitoring health unit for respiratory diseases (USMER), a sample of 375 clinics and hospitals which is part of the system of Epidemiological Surveillance implemented by the Mexican government to monitor the incidence of infectious respiratory diseases.

We use these data to estimate fatality risks associated to COVID-19 using survival analysis methods. First, we use the life-table method (Chiang 1984, Miller 1981) to estimate the cumulative proportion of COVID-19 related deaths at different durations since the onset of symptoms. We define a duration of 35 days since the onset of symptoms to obtain an estimate of the overall fatality risk.

Second, we estimate a piecewise exponential proportional hazards model (PH model) (Blossfeld et al 2019) to estimate the independent effects of demographic characteristics, comorbidities, and type of health institution on the fatality hazard associated to COVID-19. We introduce time segments corresponding to weekly time periods since the onset of symptoms, from week one to five. The PH model also includes a Gamma frailty term to control for individual heterogeneity and reduce the possible bias arising for an incomplete model specification due to unobserved variables (Paik et al 1994, Gutierrez 2002).

## Results

Table 1 presents the descriptive statistics of COVID-19 patients. The median age is 46 and 58% are males. The three most frequent comorbidities are high blood pressure (20.6%), obesity (19.4%) and diabetes (16.7%). Other comorbidities, such as asthma, cardiovascular diseases, COPD, immune suppression, and chronic liver disease, are also frequent among patients, with a prevalence between 2% and 10%. Overall, only half of patients do not present comorbidities, 27.6% have one comorbidity, 14.1% two comorbidities, and 8% three or more.

**Table 1.** Descriptive Statistics of Patients Positive to COVID-19, Mexico.

| **Variable** | **%** |  | **%** |
| --- | --- | --- | --- |
| **Age (median)** | 46 | **Prevalence of Comorbidities** |  |
| **Sex** |  | *Hypertension* | 20.6 |
| *Female* | 42.1 | *Obesity* | 19.4 |
| *Male* | 57.9 | *Diabetes* | 16.7 |
| **Source** |  | *Smoking* | 9.4 |
| *USMER* | 47.8 | *Other* | 4.7 |
| *Non-USMER* | 52.2 | *Asthma* | 3.6 |
| **Institution** |  | *Cardiovascular* | 3.0 |
| *State/Municipal* | 1.4 | *COPD* | 2.7 |
| *IMSS* | 30.7 | *Immunosup* | 2.1 |
| *IMSS-BIENESTAR, SSA* | 49.2 | *CKD* | 1.9 |
| *ISSSTE* | 6.8 | **Cumulative comorbidities** |  |
| *Private* | 10.9 | *None* | 50.3 |
| *University* | 0.9 | *One* | 27.6 |
|  |  | *Two* | 14.1 |
|  |  | *Three or more* | 8.0 |

Source: own estimates based on "Datos abiertos COVID-19", Secretaría de Salud, April 18, 2020.

Almost half of patients (47.8%) come from medical units integrated to the sentinel surveillance model sample (USMER); the other 52.2% were attended by other medical units. The distribution according to the type of medical institution shows that half of patients (49.2%) received attention by IMSS-Bienestar or the Ministry of Health, 30.7% from IMSS, 10.9% from private clinics or hospitals and 6.8% by ISSSTE. Other type of services account for only a small fraction of cases.

Figure 1 presents the cumulative proportion of deaths for patients of COVID-19, according to the number of days elapsed since the onset of symptoms. These curves are estimated using the life-table method with a correction for right-censorship. The general curve shows a pattern of acceleration during weeks 2-3 and then a decrease after 21 days.

**Figure 1.**
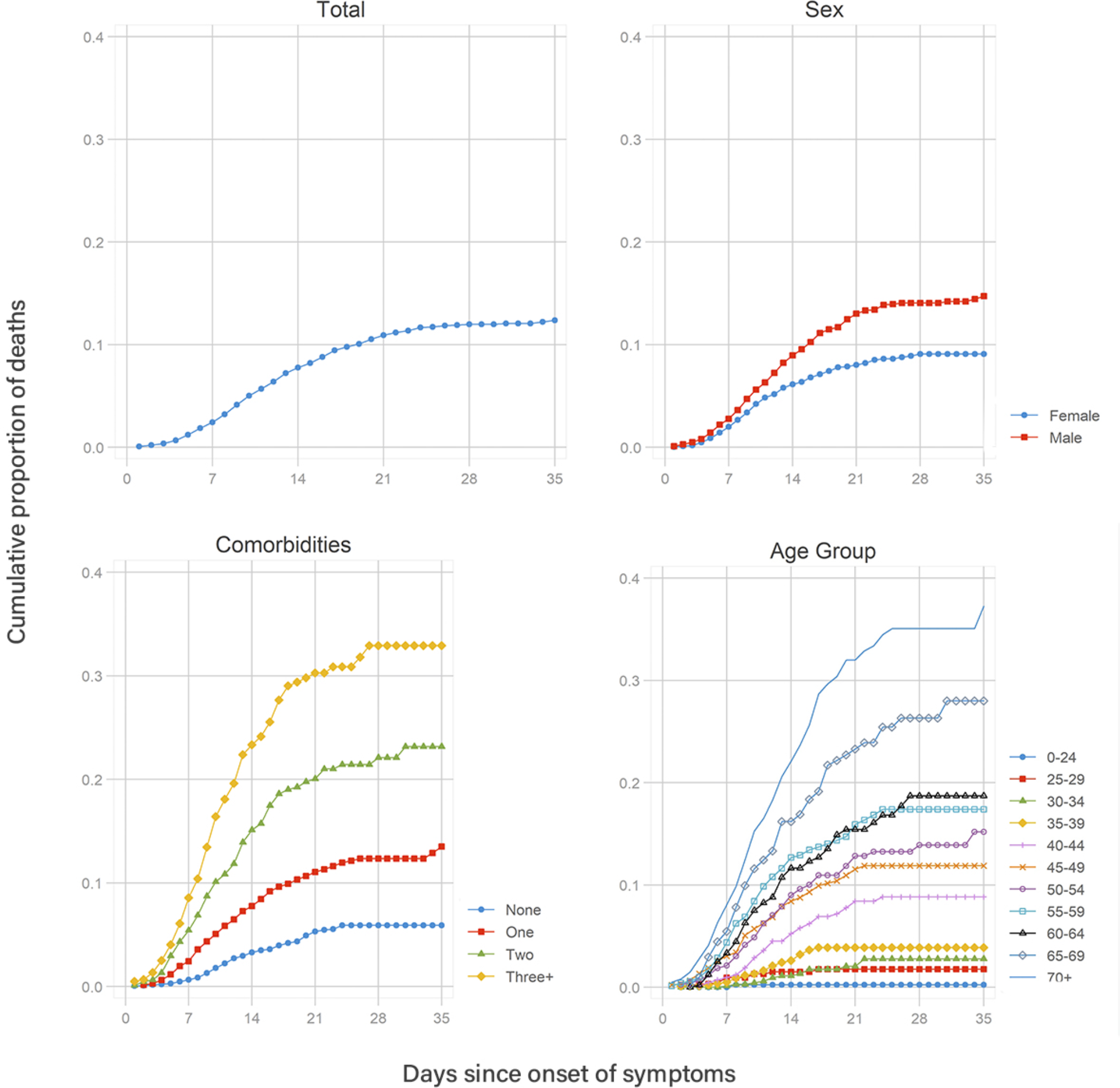
Cumulative Proportion of COVID-19 Related Deaths by Days Since Onset of Symptoms, by Sex, Number of Comorbidities and Age Group. Source: own estimates based on “Datos abiertos COVID-19”, Secretaría de Salud, April 18, 2020

The estimated death risk after 35 days (Table 2) is 12.38% (95% CI: 11.37-13.47). The risk is higher for males (14.72%, 95% CI: 13.27-16.31) than females (9.09%, 95% CI: 7.86-10.49). The risk also increases sharply with the presence of comorbidities. For patients with no comorbidities, the estimated death risk after 35 days is 5.89%, for patients with one comorbidity is 13.51%, with two comorbidities is 23.17%, and with three or more comorbidities is 32.93%. There is also a positive association with age: patients with less than 25 years of age have an estimated death risk of 0.24%, those with ages 40-44 a risk of 8.83% and those with 70+ years a risk of 37.27%.

**Table 2.**
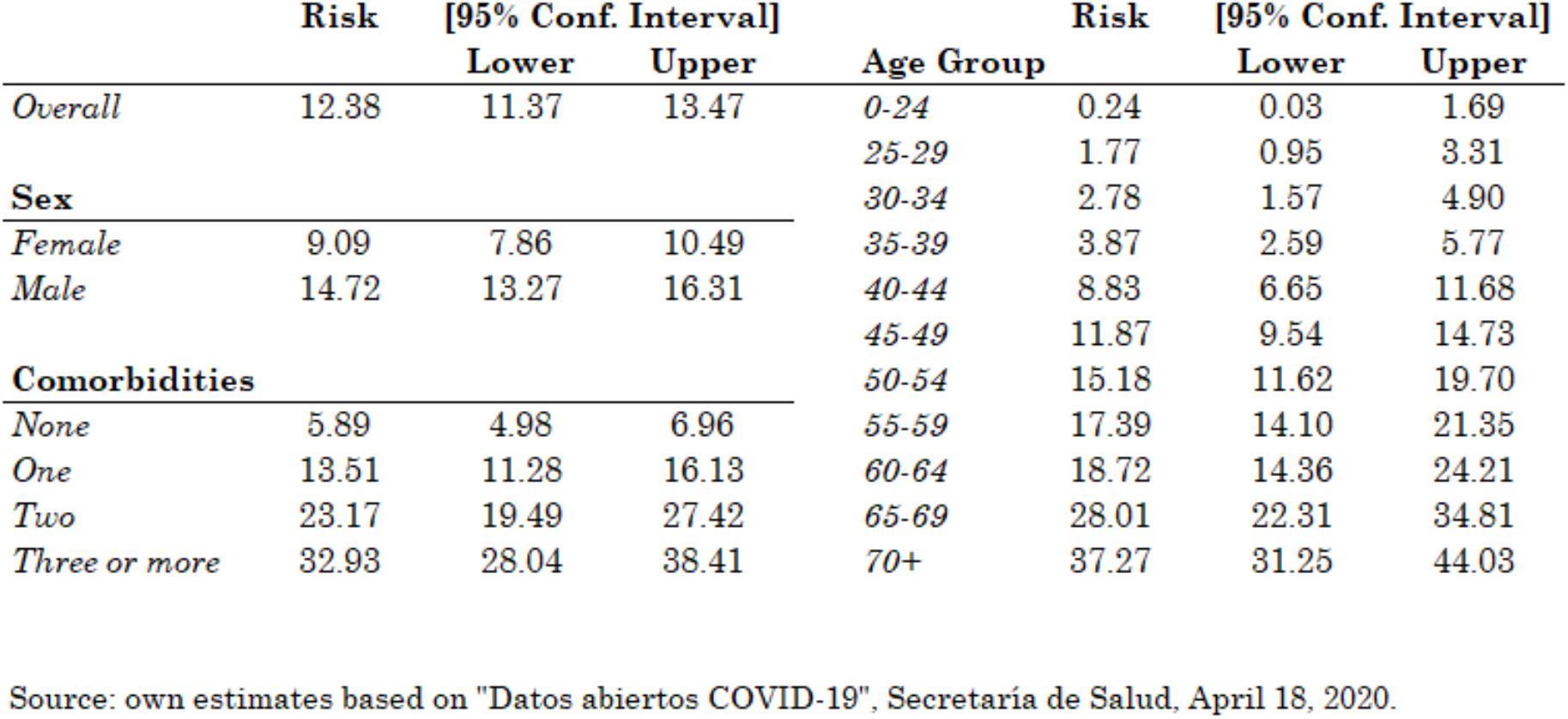
Estimation Fatality Risk of COVID-19 Patients After 35 Days of Onest of Symptoms.

These estimates are based on bivariate associations, so they may reflect the effects of correlated factors not controlled in the estimation. In order to evaluate the independent effect of each factor we estimated a piecewise exponential hazard model for the risk of a COVID-19 related death (Table 3). As explained in the methodological section, this model includes a gamma term for unobserved heterogeneity, which may help to reduce the biases associated to unobserved variables.

**Table 3.** Results from Piecewise Exponential Model for the Risk of Death Among Symptomatic Confirmed Patients of COVID-19, Mexico.

| Variable | Hazard Ratio | P-Value | [95% CI]<br>Lower Upper |  | Variable | Hazard Ratio | P-Value | [95% CI]<br>Lower Upper |  |
| --- | --- | --- | --- | --- | --- | --- | --- | --- | --- |
| **Male** | 1.589 | 0.000 | 1.329 | 1.899 | **Comorbidities** |  |  |  |  |
| **Non-USMER** | 0.644 | 0.000 | 0.544 | 0.763 | *Hypertension* | 1.378 | 0.008 | 1.086 | 1.749 |
| **Week** |  |  |  |  | *Obesity* | 1.777 | 0.000 | 1.415 | 2.231 |
| *First* | 1.000 | - | - | - | *Cardiovascular* | 0.872 | 0.449 | 0.611 | 1.244 |
| *Second* | 1.842 | 0.000 | 1.510 | 2.249 | *Diabetes* | 1.730 | 0.000 | 1.360 | 2.202 |
| *Third* | 1.314 | 0.065 | 0.983 | 1.756 | *Immunosup* | 1.696 | 0.008 | 1.148 | 2.505 |
| *Fourth* | 0.473 | 0.008 | 0.273 | 0.821 | *COPD* | 1.770 | 0.001 | 1.269 | 2.469 |
| *Fifth* | 0.253 | 0.020 | 0.079 | 0.809 | *Asthma* | 0.776 | 0.319 | 0.471 | 1.278 |
| **Age Group** |  |  |  |  | *CKD* | 2.679 | 0.000 | 1.789 | 4.014 |
| *0-24* | 0.038 | 0.001 | 0.005 | 0.270 | *Smoking* | 1.049 | 0.738 | 0.793 | 1.388 |
| *25-29* | 0.202 | 0.000 | 0.104 | 0.392 | *Other* | 1.001 | 0.995 | 0.684 | 1.464 |
| *30-34* | 0.218 | 0.000 | 0.121 | 0.394 | *None or one* | 1.000 | - | - | - |
| *35-39* | 0.357 | 0.000 | 0.225 | 0.566 | *Two* | 1.072 | 0.648 | 0.796 | 1.444 |
| *40-44* | 0.756 | 0.122 | 0.530 | 1.078 | *Three or more* | 0.747 | 0.263 | 0.449 | 1.244 |
| *45-49* | 1.000 | - | - | - | **Institution** |  |  |  |  |
| *50-54* | 1.171 | 0.328 | 0.854 | 1.605 | *State/Municipal* | 1.000 | - | - | - |
| *55-59* | 1.411 | 0.030 | 1.035 | 1.923 | *IMSS* | 1.553 | 0.188 | 0.807 | 2.988 |
| *60-64* | 1.304 | 0.128 | 0.926 | 1.836 | *IMSS-BIENESTAR, SSA* | 0.887 | 0.719 | 0.463 | 1.700 |
| *65-69* | 2.016 | 0.000 | 1.458 | 2.787 | *ISSSTE* | 0.821 | 0.583 | 0.406 | 1.660 |
| *70+* | 2.608 | 0.000 | 1.931 | 3.522 | *Private* | 0.398 | 0.014 | 0.191 | 0.832 |
|  |  |  |  |  | *University* | 1.201 | 0.686 | 0.493 | 2.923 |
| *theta* | 0.086 |  |  |  | *LR chibar2(01)* | 0.05 |  |  |  |
|  |  |  |  |  | *Prob >= chibar2* | 0.408 |  |  |  |
| *Subjects (N)* | 7,388 |  |  |  |  |  |  |  |  |
| *Failures* | 639 |  |  |  | *LR chi2(33)* | 934.34 |  |  |  |
| *Log likelihood* | -2183.87 |  |  |  | *Prob > chi2* | 0.000 |  |  |  |

Source: own estimates based on "Datos abiertos COVID-19", Secretaría de Salud, April 18, 2020.

The hazard ratios confirm the time-dependence pattern observed in the descriptive analysis. Compared to the first week of symptoms, the estimated hazard rate increases 84% in the second week and 31% in the third week, and then reduces in half (HR 0.473) in the fourth week and almost 75% (HR 0.253) in the fifth week.

The higher risk of a COVID-19 related fatality among confirmed male patients is also corroborated by the model. Even after controlling by age and comorbidities, the estimated death hazard is 59% higher for males (HR=1.589, 95% CI 329-899). The effects of age are also positive and statistically significant. Compared to a patient between 45-49 years of age, the estimated hazard of death is 161% higher for a patient with 70+ years (HR=2.608, 95% CI 1.931-3.522) and 96% lower for a patient under 25 years (HR=0.038, 95% CI 0.005-0.270).

Not all pre-existing comorbidities affect the hazard of a COVID-19 related death. The model does not identify significant effects for asthma, cardiovascular disease, smoking and “other” comorbidities. In contrast, high blood pressure increases the estimated hazard by 38% (HR=1.378, 95% CI: 1.086-1.749), immune suppression by 70% (HR=1.696, 95% CI: 1.148-2.505), diabetes by 73% (HR=1.730, 95% CI: 36.0-120.2), COPD by 77% (HR=1.770, 95% CI: 1.269-2.469), obesity by 78% (HR=1.776, 95% CI: 1.415-2.231), and chronic liver disease by 168% (HR=2.679, 95% CI: 1.789-4.014). There are no significant interaction effects associated to the presence of two or more of these pre-existing conditions.

Finally, the estimated fatality hazard is 36% lower for patients who received attention in non-USMED units (HR=0.644, 95% CI: 0.544-0.763). This may reflect a negative selectivity of patients who receive attention in USMED units, which are generally more equipped than Non-USMED units and therefore may concentrate the most severe cases. This selectivity may also explain the higher fatality risk for patients attended by IMSS, compared to IMSS-BIENESTAR/SSA, ISSSTE, and private institutions. However, differences in the quality and procedures of attention between institutions cannot be discarded, particularly to explain the larger reduction in the fatality hazard in private institutions (HR=0.257, 95% CI: 0.170-0.386).

## Conclusions

In this paper we apply survival analysis methods to estimate the COVID-19 fatality risk and associated risk factors for symptomatic confirmed patients of COVID-19 in Mexico, using individual-level data recently released by the Ministry of Health.

The fatality risk after 35 days of onset of symptoms estimated using the life-table method is 12.38%, a figure considerably higher than the 8.67% obtained by the simple ratio of deaths vs. confirmed cases. This difference is mostly explained by the right-censorship in cross-sectional data: a considerable proportion of confirmed patients are still at risk of death and some of them eventually will die, but they are not currently considered in the calculation of the cross-sectional fatality rate. The fatality risk estimated with the life-table method introduces corrections for this bias, and therefore may be closer to the final fatality risk for symptomatic confirmed patients of COVID-19 in Mexico.

The estimated fatality hazard for symptomatic confirmed patients of COVID-19 is higher during the second week of the onset of symptoms. It is also higher for males and sharply increases with age. The preexistence of certain comorbidities, such as chronic kidney disease, COPD, obesity, diabetes, immune suppression, and hypertension is also a risk factor which significantly increases the fatality hazard. There are no significant effects associated to cardiovascular disease, asthma, and smoking.

These results must be taken with caution because they represent only a subset of COVID-19 cases, those with symptoms serious enough to seek medical attention and who tested positive for the presence of the virus. Mexico has adopted a policy of restriction of tests to such cases. Compared to other countries with significant number of COVID-19 cases, Mexico has one of the lowest number of tests per thousand (0.28, *versus* a median of 6.28 per thousand for a group of 50 countries with updated information up to April 18)^2^, and therefore the selectivity of confirmed cases among those with serious symptoms is likely higher than in most countries. This selectivity, along with the high prevalence of comorbidities among the adult population, might explain the high fatality risk observed in Mexico.

In sum, our results may be best interpreted as reflecting the fatality risks for patients with serious symptoms who seek medical attention in Mexico, and caution must be exercised to extrapolate them to the overall infected population or to other countries with different epidemiological profiles or COVID-19 testing policies.

## Data Availability

All data used in this paper is available to the public in the provided URL.

https://www.gob.mx/salud/documentos/datos-abiertos-152127

## Footnotes

1 https://www.gob.mx/salud/documentos/datos-abiertos-152127

2 https://ourworldindata.org/covid-testing

## Notes

### Competing Interest Statement

The authors have declared no competing interest.

### Funding Statement

No external funding was received.

